# The Role of Neighborhood Socioeconomic Environment in the Association Between Glycemic Control and the Developing Brain

**DOI:** 10.64898/2026.03.31.26349868

**Authors:** Anisha Chandra, Eustace Hsu, Shan Luo

## Abstract

**Objective:** To investigate overall and neighborhood socioeconomic deprivation moderated associations between glycemic control and brain structure in youth.

**Research Design and Methods:** This was a cross-sectional study of 705 healthy 11–12-year-olds across 21 study sites in the United States. Data was obtained from the Adolescent Brain and Cognitive Development (ABCD) Study®. Glycemic control was assessed using hemoglobin A1c (HbA1c), brain structure was evaluated via MRI, and neighborhood deprivation was measured with the Area Deprivation Index (ADI). Mixed effects models were used to examine relationships between HbA1c, brain structure and ADI controlling for sociodemographic covariates. Stratified analysis was performed by tertiles of ADI.

**Results:** Higher HbA1c was associated with lower mean cortical thickness (CT) and smaller total cortical gray matter volume (GMV). One percent increase in HbA1c corresponded to a 0.024 mm reduction in mean CT and a 9,611 mm³ reduction in total cortical GMV. Regionally, higher HbA1c was associated with thinner cortex and smaller gray matter volumes primarily in the frontal, cingulate and occipital areas. There was a significant interaction of HbA1c and ADI on total GMV, which was driven by significant negative associations of HbA1c with total GMV in the high ADI group, and medium ADI group, but not the low ADI group.

**Conclusions:** Mild elevations in HbA1c, even within the non-diabetic range, are linked to early brain structural changes, particularly in youth from neighborhoods with greater socioeconomic deprivation. These results highlight the interplay between metabolic health and neighborhood deprivation on shaping brain development in youth.

## Introduction

Obesity and type 2 diabetes (T2D) rates have continued to rise over the past several decades in adults as well as adolescents. Approximately one in five children in the US are overweight or obese, predisposing them to diabetes (1). The estimated prevalence of T2D in youth nearly doubled between 2001 and 2017, with higher incidence and T2D related complications and comorbidities in youth from lower socioeconomic backgrounds (2). Compared to previous generations, the current generation of youth is more overweight and T2D is being diagnosed at earlier ages, affirming that these conditions are key public health priorities (3). T2D in children comes with several potential complications, including neurological issues.

The developing brain is highly reliant on glucose for fuel and growth. Functional imaging studies have demonstrated a network of brain regions sensitive to *acute* changes in the circulating levels of glucose, including the hypothalamus, limbic regions, and the prefrontal cortex (4). It is less known how *chronic* exposure to high levels of glucose impacts the developing brain. Structural imaging studies have found that youth with T2D had smaller global and regional gray matter volumes, primarily in the prefrontal and limbic regions (e.g., striatum, hippocampus, amygdala, thalamus) (4–7). Such structural abnormalities may manifest as reduced cognitive function, a finding observed in youth with T2D (7,8). Moreover, higher HbA1c was associated with smaller prefrontal volumes and greater cerebral atrophy in youth with T2D, suggesting that poor glycemic control may contribute to these observed structural brain abnormalities in youth with T2D (5).

While prior studies have demonstrated brain structure impairments in youth with T2D, it remains unknown whether the changes in the brain occur before onset of diabetes or consequences of diabetes. Furthermore, existing studies have not shown how individual variabilities in HbA1c relate to brain structure in healthy controls, where early alterations may indicate vulnerability before diabetes develops. Thus, it remains unknown the impact of glucose control on the structural development of the brain in typically developing youth prior to onset of T2D.

Early brain changes linked to glucose regulation may not occur in isolation but within the broader context of socioeconomic disadvantage. A growing body of evidence shows that higher neighborhood socioeconomic deprivation was associated with poorer glycemic control in youth with diabetes, and lower global and regional white and gray matter volumes, cortical thickness, surface area, and subcortical volumes (9–11). While several studies have highlighted racial-ethnic disparities underlying T2D in youth, it remains unknown whether different neighborhoods have differential relationships between HbA1c and the brain.

The purpose of our study was to evaluate the relationship between glycemic control and youth brain development, specifically how neighborhood socioeconomic environment moderates these relationships. Given prior findings in adults and children, we hypothesized that youth with higher HbA1c would have smaller structural brain measures, particularly smaller volumes in the prefrontal and limbic regions. Neighborhood socioeconomic deprivation may moderate these associations, such that there would be a stronger negative relationship between HbA1c and structural brain measures in more socioeconomically deprived neighborhoods.

## Research Design and Methods

### Participants

The ABCD® Study is a 10-year longitudinal study of adolescent development. Participants aged 9-10 years old were initially recruited from 21 study sites across the U.S. between September 2016 and August 2018. Parents completed written informed consent and children provided assent for study participation. Study procedures were approved by the centralized Institutional Review Board (IRB) at the University of California, San Diego (IRB #160091), in addition to each study site with its respective IRB. Further details for the ABCD Study, including design, recruitment, and screening criteria, have been reported previously (12). For the current analysis, data were obtained from the ABCD® 4.0 data release (13), and focused on two-year follow-up between 3/3/2019 and 3/17/2020, when imaging and HbA1c data were collected.

Participants were excluded if they met the following criteria: not fluent in English, history of seizures, birth more than 12 weeks premature, birth weight less than 1200 grams, complications at birth, substance use disorder, intellectual disability, traumatic brain injury, major neurological disorders, schizophrenia, and other medical conditions considered exclusionary (12). Further exclusion criteria were applied based on availability and validity of blood samples and neuroimaging data, as well as covariates described in the following sections. In summary, our final sample (N=705) included participants with quality-controlled neuroimaging measurements and complete data for all variables used in analyses (Supplemental Fig. S1).

### HbA1c

Blood samples were collected and analyzed for HbA1c level. Participants were excluded with a parental-reported diabetes diagnosis at any time-point in the study, or with HbA1c of at least 6.5 (*n*=9).

### Neuroimaging

Methods for MRI data collection were optimized and harmonized for 3-Tesla scanners across multiple platforms (General Electric MR 750, Philips Achieva dStream and Ingenia, and Siemens Prisma and Prisma Fit) across 21 ABCD® study sites (14,15). Structural images were collected with T1-weighted anatomical scans. Cortical surface reconstruction and subcortical segmentation was completed at the ABCD® Data Analytics and Informatics Center (DAIC) via FreeSurfer (version 5.3.0), producing estimates of global and regional cortical thickness (mm), cortical surface area (mm^2^), and cortical and subcortical gray matter, and white matter volume (mm^3^).

Quality control (QC) was conducted to evaluate each cortical surface reconstruction for five categories of inaccuracy: severity of motion, intensity inhomogeneity, white matter underestimation, pial overestimation, and magnetic susceptibility artifact. Images were excluded for failure to pass all categories of QC, as well as for abnormal radiological findings. Additionally, the top and bottom 1% of each neuroimaging measure was winsorized for outlier detection.

### Imaging Data Harmonization

Neuroimaging data was harmonized using the ComBat method, an empirical Bayes-based method for harmonizing data across multiple scanners, with the ability to remove additive and multiplicative effects related to the scanner (16,17). Harmonization was processed using the unique scanner identification as the “batch” effect, and sex, interview age, and HbA1c as biological covariates. In this way, the scanner-related variances between sites were removed while the biological variances across sites were retained.

### Area Deprivation Index

Primary address was reported by the caregiver and matched with the 2010 US census tracts to identify neighborhoods. Area Deprivation Index (ADI) was calculated for individual census tracts based on 17 characteristics derived from the American Community Survey (2011-2015) (18). ADI is a comprehensive measurement of neighborhood socioeconomic deprivation, with higher ADI scores representing greater neighborhood deprivation (19). National percentiles of ADI were obtained by ranking across all census tracts, to obtain an ADI percentile score corresponding to each participant’s residential history. ADI percentile was grouped into tertiles for ease of interpretation.

### Covariates

Sex was categorized as the child’s sex at birth, based on caregiver reports. Age was the child’s age in months at the time point of study visit. Race/ethnicity was categorized as Hispanic, non-Hispanic White, non-Hispanic Black, non-Hispanic Asian, and Other. Parental education was modeled as a binary variable indicating whether at least one parent has obtained a bachelor’s degree. Yearly household income was quantified in three categories: less than $50,000, between $50,000 and $99,999, and at least $100,000. Pubertal stage was calculated using the parent-reported pubertal development scale.

### Data Analysis

Linear mixed effects models were conducted to examine associations between HbA1c (independent variable) and global brain measurements (dependent variables), adjusting for covariates with impact on brain structure. Global brain measurements included total cortical surface area, mean cortical thickness (CT), total cortical gray matter volume (GMV), subcortical GMV, and cerebral white matter volume. Family ID was included in the model as random effects to account for shared family membership; furthermore, age, sex, race/ethnicity, parental education, household income, pubertal stage, and handedness were included as covariates. Intracranial volume (ICV) was included as an additional covariate in models of volumetric measures.

Subsequent region-of-interest (ROI) analyses were completed to follow up with significant associations of HbA1c with global brain measures. Cortical GMV and mean CT of 34 ROIs defined bilaterally on the Desikan-Killiany atlas were included. Correlations in each ROI were assessed using models predicting bilateral brain structure based on HbA1c level by hemisphere (right, left) interaction. Family ID was included in the model as random effects; furthermore, subject identification was included as a random effect to pool across hemispheres. Models used the following covariates: age, sex, race/ethnicity, parental education, household income, pubertal stage, and handedness. Models of GMV ROIs included ICV as an additional covariate.

To further examine the impact of neighborhood socioeconomic deprivation on the relationships between HbA1c and brain, interaction analysis of HbA1c x ADI (tertile) was performed on global and regional brain measurements identified as significant in the whole cohort model. Family ID was modeled as a random effect. Age, sex, race/ethnicity, pubertal stage, and handedness were included as covariates, with ICV as an added covariate for volumetric models.

Linear mixed effects models were fitted in R with the *lme4* package. Standardized betas were reported, representing the standard-deviation increase in the dependent variable corresponding to a one standard-deviation increase in HbA1c. 95% Wald confidence intervals were calculated based on the local curvature of the likelihood surface. *P*-values were calculated using Satterthwaite’s method in the *lmerTest* package. Tests of significance (2-tailed) were corrected for multiple comparisons using the Benjamini-Hochberg false discovery rate (FDR) correction, with *P*< 0.05 as the corrected threshold for significance. Relationships of regional brain measures with HbA1c, pooled across hemisphere, were reported using the *emmeans* function in the *emtrends* package.

### Data and Resource Availability

Data used in the preparation of this article were obtained from the Adolescent Brain Cognitive Development (ABCD) Study (https://abcdstudy.org), held in the National Institute of Mental Health (NIMH) Data Archive (NDA). This is a multisite, longitudinal study designed to recruit more than 10,000 children aged 9 to 10 years and follow them over 10 years into early adulthood.

## Results

### Demographics

Descriptions of the sample are shown in **Table 1**. The average age of participants was 11.9 years with a range between 10 years 7 months, and 13 years 2 months. 43.3% of participants were female, 16.6% of participants were Hispanic or Latino, 60.7% non-Hispanic White, 10.4% non-Hispanic Black, 1.3% Asian, and 11.1% Other race. 21.7% of participants lived in households with less than $50,000/yr income, 28.1% with between $50,000/yr and $99,999/yr, and 50.2% above $100,000/yr. 65.0% of participants had at least one parent who graduated college. 22.8% of participants were pre-pubertal, 26.4% were in early puberty, 33.3% were in mid-puberty, 16.6% were in late puberty, and 0.9% were post-puberty. HbA1c was correlated with race/ethnicity (*F*_4,514_ = 9.68, *P*<0.001) and parental education (*F*_1,567_=13.99, *P*<0.001).

**Table 1.**
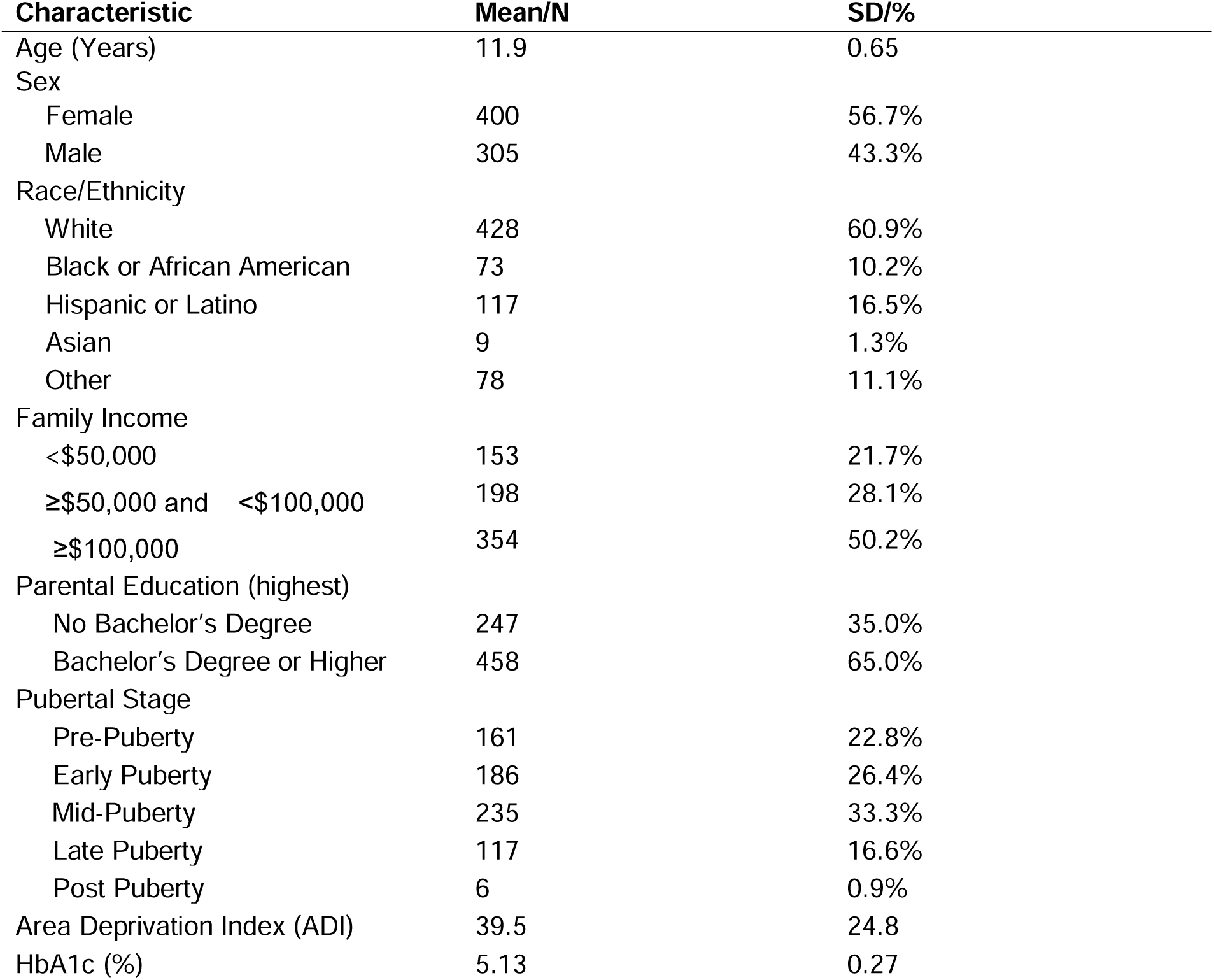
Participant Demographics.

### HbA1c and Global and Regional Brain measures

Higher HbA1c was associated with lower mean CT (β (95% CI) = -0.090 (-0.164, -0.015), FDR corrected *P*=0.046) and smaller total cortical GMV (β (95% CI) = -0.048 (-0.085, -0.010), FDR corrected *P*=0.046) (**Fig. 1**). One percent increase in HbA1c was associated with a -0.024 mm decrease in mean CT and 9,611 mm^3^ decrease in total GMV. HbA1c was not significantly associated with total cortical surface area, total subcortical GMV, and cerebral white matter volume (**Table 2**).

**Figure 1.**
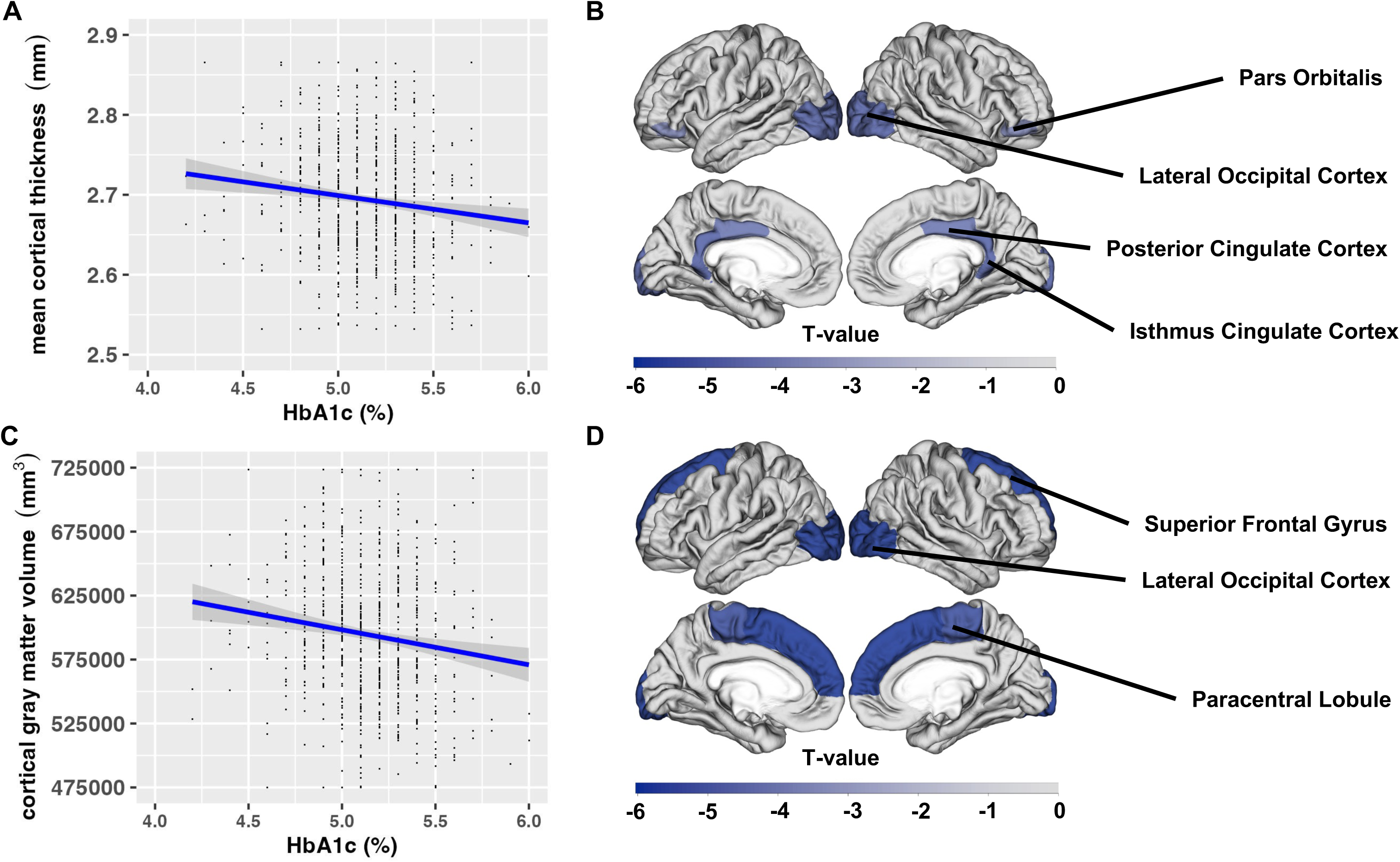
Association of hemoglobin A1c (HbA1c) levels (%) with global and regional brain measurements. **A).** Scatterplot displays the relationship of HbA1c % with mean cortical thickness (CT). One percent increase in HbA1c was associated with a 24 mm or 0.9% decrease in mean CT. **B).** T value denotes regions significantly associated with GDM exposure from linear mixed effects models (FDR corrected P<0.05), where family ID nested within scanner ID, as well as subject, were included as random effects, and handedness, intracranial volume, age, sex, pubertal status, race/ethnicity, family income, and parental education were included as fixed effected. All regions of interest analysis were bilateral in the models. **C).** Scatterplot displays the relationship of HbA1c % with total cortical gray matter volume (GMV). One percent increase in HbA1c was associated with a 9,611 mm³ or 1.6% decrease in total cortical GMV. **D).** Associations between HbA1c and child regional cortical thickness in significant regions of interest. T value denotes regions significantly associated with GDM exposure from linear mixed effects models (FDR corrected *P*<0.05), where family ID nested within scanner ID, as well as subject, were included as random effects, and handedness, age, sex, pubertal status, race/ethnicity, family income, and parental education were included as fixed effects. All regions of interest analysis were bilateral in the models.

**Table 2.** Associations between HbA1c and Global Brain Measurements.

| Global Brain Measurements | $\beta$ (95% CI)* | FDR-adjusted<br>*P*-value† |
| --- | --- | --- |
| Total Cortical Surface Area | -0.009 (-0.069, 0.052) | 0.782 |
| Mean Cortical Thickness | -0.090 (-0.164, -0.015) | **0.046** |
| Cortical Gray Matter Volume | -0.048 (-0.085, -0.010) | **0.046** |
| Subcortical Gray Matter Volume | -0.028 (-0.069, 0.013) | 0.306 |
| Cerebral White Matter Volume | 0.010 (-0.027, 0.047) | 0.753 |

\* Standardized $\beta$ (95% CI) from linear mixed effects models. Positive $\beta$ indicates higher HbA1c is associated with larger brain measurement, negative $\beta$ indicates the opposite relationship
† Multiple comparisons are conducted with Benjamini-Hochberg FDR correction. Boldface indicates significance at the corrected threshold of $P < 0.05$ .

ROI analysis of GMV found that higher HbA1c was associated with smaller GMV in bilateral lateral occipital cortex (β (95% CI) = -0.093 (-0.148, -0.039), FDR corrected *P*=0.025), paracentral lobule (β (95% CI) = -0.079 (-0.132, -0.026), FDR corrected *P*=0.040), and superior frontal gyrus (β (95% CI) = -0.078 (-0.127, -0.030), FDR corrected *P*=0.027) (Supplemental Table S1, **Fig. 1**). A one percent increase in HbA1c was associated with 1402 mm^3^ decreases in volume of the lateral occipital cortex, 374 mm^3^ in the paracentral lobule, a 282 mm^3^ in the posterior cingulate cortex, and 1976 mm^3^ in the superior frontal cortex.

ROI analysis of CT found greater HbA1c was correlated with thinner isthmus cingulate cortex (β (95% CI) = -0.107 (-0.172=, -0.043), FDR corrected *P*=0.018), thinner lateral occipital cortex (β (95% CI) = -0.101 (-0.170, -0.033), FDR corrected *P*=0.026), thinner pars orbitalis (β (95% CI) = -0.093 (-0.157, -0.029), FDR corrected *P*=0.037), and posterior cingulate cortex (β (95% CI) = -0.107 (-0.171, -0.044), FDR corrected *P*=0.018) (Supplemental Table S2, **Fig. 1**). A one percent increase in HbA1c was associated with 0.054 mm decreases in mean thickness of isthmus cingulate, 0.040 mm in lateral occipital cortex, 0.055 mm in pars orbitalis, and 0.043 in posterior cingulate cortex.

### HbA1c and Brain, moderated by ADI

There was a significant interaction of HbA1c and ADI on total GMV (*F*_2,642_=35.235, FDR corrected *P*<0.001), which was driven by significant associations of HbA1c with total GMV in the medium ADI group (β (95% CI) = -0.087 (-0.158, -0.015), FDR corrected *P*=0.036), and the high ADI group (β (95% CI) = -0.071 (-0.133, -0.010), FDR corrected *P*=0.024), but not the low ADI group (β (95% CI) = -0.004 (-0.067, 0.059), FDR corrected *P*=0.897) (**Fig. 2**).

**Figure 2.**
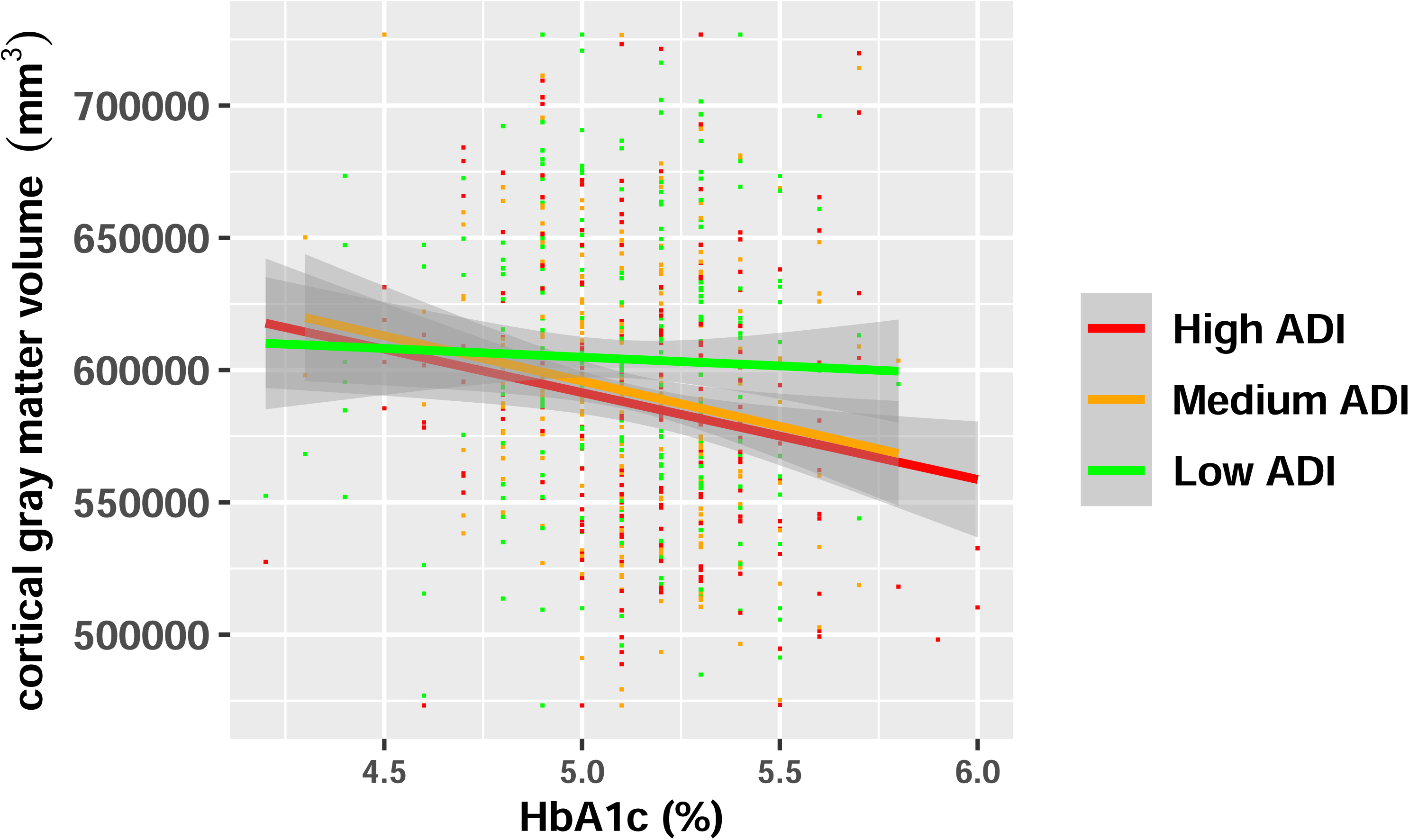
Relationships between HbA1c levels (%) and adjusted total cortical gray matter volume (GMV) in children from neighborhoods with low, medium, and high area deprivation index (ADI). Lines represent interaction model regression fits and shaded areas around the fitted lines represent standard error of the fits. This shows a significant interaction of HbA1c and ADI on total GMV.

Interaction of HbA1c and ADI was not significant for mean CT (*F*_2,648_=2.133, FDR corrected *P*=0.119). Associations of HbA1c with mean CT was significant in the high ADI group (β (95% CI) = -0.156 (-0.278, -0.034), FDR corrected *P*=0.024), but not for the medium ADI (β (95% CI) = -0.121 (-0.264, 0.021), FDR corrected *P*=0.095), nor low ADI group (β (95% CI) = -0.004 (-0.067, 0.059), FDR corrected *P*=0.897).

Interaction of HbA1c and ADI was not observed in regional GMV in the lateral occipital cortex (*F*_2,653_=0.537, FDR corrected *P*=0.594), paracentral lobule (*F*_2,649_=0.521, FDR corrected *P*=0.594), and superior frontal gyrus (*F*_2,646_=1.535, FDR corrected *P*=0.216). There were no significant interactions of HbA1c and ADI in regional CT in the isthmus cingulate (*F*_2,654_=0.600, FDR corrected *P*=0.549), lateral occipital (*F*_2,651_=1.834, FDR corrected *P*=0.373), pars orbitalis (*F*_2,648_=0.892, FDR corrected *P*=0.547), and posterior cingulate cortex (*F*_2,650_=0.684, FDR corrected *P*=0.373). Detailed results in each ADI tertile can be found in Supplemental Tables S3 and S4.

## Discussion

The aim of our study is twofold: first, to examine the relationships between glycemic control and brain structure development in healthy developing youth without diabetes; second, to understand the role of neighborhood socioeconomic deprivation in moderating these relationships. Our findings revealed that higher HbA1c levels, which may indicate worse glycemic control, were associated with lower mean CT and smaller total cortical GMV in youth without diabetes. Notably, the negative associations between HbA1c and GMV were only significant in youth living in neighborhoods with higher socioeconomic deprivation. These results suggest that subtle variations in glycemic control, even within non-diabetic range, may influence pediatric brain development. Moreover, these results highlight the potential amplifying effect of neighborhood socioeconomic deprivation and provide support for diabetes prevention efforts targeting youth from disadvantaged neighborhoods before onset of diabetes.

Our results are consistent with prior studies that demonstrated lower GMVs in youth with chronic exposure to high levels of glucose (20–22). Since our study included a sample of youth without diabetes, it presents novel findings in a population that has not been studied extensively, and our study results suggest that subtle dysregulation in glycemic control may be associated with early signs of brain atrophy detectable *before* the onset of diabetes. Our primary finding that higher HbA1c levels are associated with smaller brain volumes supports previous research on the detrimental impact of poor glycemic control on brain regions such as the prefrontal cortex and limbic areas (23,24). The ROI analysis found that some of the effects are specific to particular cortices which is consistent with the notion that chronically elevated glucose levels can disrupt the normal development of brain regions responsible for executive functioning, emotional regulation, and cognitive processing (25–29).

We observed that a 1% increase in HbA1c was linked to a 1.6% decrease in total cortical GMV and 0.9% decrease in mean CT, which could suggest that chronic exposure to elevated glucose may hinder neurodevelopment. Multiple studies have shown that decreases in total cortical GMV and mean CT can have clinical implications in various neurological and psychiatric disorders, such as dementia, schizophrenia, and depression in adults (30–33). A recent study by Shin et al. showed no significant relationship between HbA1c and mean CT in healthy adults, but a negative relationship between HbA1c and mean CT only in adults with prediabetes and diabetes (34). Our study challenges this finding, as there is a negative relationship between HbA1c and mean CT in healthy developing youth. This indicates that higher HbA1c may have a stronger impact on cortical development, as the adolescent brain is particularly sensitive to environmental influences. However, there is no established threshold for cortical GMV and CT that implies clinical significance, as these are only a couple of factors among a variety of influences on neurodevelopment.

In addition, our study is one of the first to examine the role of neighborhood socioeconomic deprivation as a moderator of the glycemic control-brain relationship in youth without diabetes, extending previous research that has primarily focused on individuals with diabetes. This novel approach highlights the importance of early environmental factors, such as neighborhood deprivation, in shaping brain development even in the absence of diabetes. These findings are consistent with the growing body of evidence suggesting that environmental stressors, including socioeconomic disadvantage, contribute to neurodevelopmental outcomes in youth (10).

The significant interaction between HbA1c and ADI on total GMV underscores the importance of considering the broader socioeconomic context when examining health disparities. In neighborhoods with higher socioeconomic deprivation, the negative relationship between glycemic control and brain structure was more pronounced. This interaction suggests that these negative effects might be exacerbated by neighborhood-related conditions like limited access to resources, higher levels of stress, and poorer overall health infrastructure (9,10). It is possible that youth in such environments face additional psychosocial and environmental stressors that may further compromise brain development, particularly in the context of metabolic disturbances like those associated with dysregulated glucose levels.

Our study has limitations that should be acknowledged. We did not observe significant interactions between HbA1c and ADI for mean CT, nor for regional GMVs in the occipital cortex and superior frontal gyrus. This could suggest that the impact of neighborhood deprivation on brain structure may be more pronounced in specific global brain measures (e.g., total GMV) rather than in more localized areas. It is also possible that the effects of neighborhood deprivation might vary across neuroanatomical regions, with some areas being more sensitive.

Furthermore, the cross-sectional design of the ABCD Study limits our ability to draw causal inferences regarding the relationship between glycemic control, neighborhood deprivation, and brain structure. Longitudinal studies will be important for establishing the sequence of these relationships and for examining whether the effects of neighborhood deprivation on brain structure are cumulative over time. Although we controlled for a range of sociodemographic factors, there may be other unmeasured confounders, such as dietary factors, physical activity or sleep patterns, that could influence both glycemic control and brain development. Lastly, the current sample included only youth without diabetes, which limits the generalizability of our findings to populations with chronic metabolic disorders.

Future research should explore whether brain markers can predict the onset of diabetes in children, particularly those from disadvantaged neighborhoods. Targeted diabetes prevention in children is crucial, but individual- and family-level efforts may not be sufficient on their own. Preventive measures should prioritize low socioeconomic populations, especially in the context of gentrification, which may unintentionally lower A1c levels but also raise concerns. Systemic solutions, such as effective urban planning to enhance neighborhood safety, walkability, and access to groceries, transportation, and green spaces, could play a significant role in improving public health outcomes. Multi-level action, including government policies promoting nutrition and physical activity, is essential to make preventive habits more accessible for vulnerable groups.

Longitudinal studies are needed to clarify the relationships between glycemic control, brain structure and diabetes risk. It is also important to examine how neighborhood characteristics, such as socioeconomic deprivation and access to food, influence A1c levels and brain health, and whether these factors operate independently or interact. Understanding these pathways can guide interventions in low socioeconomic neighborhoods and support policies that address built environments and social factors like safety, healthcare access, and stress, all of which are linked to the development of diabetes in youth.

## Conclusions

We observed that higher HbA1c levels were associated with lower cortical thickness and smaller global and regional cortical gray matter volumes, with an exaggerated relationship between these variables demonstrated in youth from disadvantaged neighborhoods. Our study provides evidence that neighborhood socioeconomic deprivation may exacerbate the negative effects of subtle elevations in HbA1c on brain structure in healthy developing youth. These results also provide preliminary evidence to support targeted diabetes interventions in youth from socioeconomically deprived neighborhood, which may help prevent metabolic diseases in vulnerable populations and reduce the burden of diabetes-related neurological complications.

## Data Availability

Data used in the preparation of this article were obtained from the Adolescent Brain Cognitive Development (ABCD) Study (https://abcdstudy.org), held in the National Institute of Mental Health (NIMH) Data Archive (NDA). This is a multisite, longitudinal study designed to recruit more than 10,000 children aged 9 to 10 years and follow them over 10 years into early adulthood. The ABCD Study is supported by the NIH and additional federal partners under award numbers U01DA041048, U01DA050989, U01DA051016, U01DA041022, U01DA051018, U01DA051037, U01DA050987, U01DA041174, U01DA041106, U01DA041117, U01DA041028, U01DA041134, U01DA050988, U01DA051039, U01DA041156, U01DA041025, U01DA041120, U01DA051038, U01DA041148, U01DA041093, U01DA041089, U24DA041123, and U24DA041147. A full list of supporters is available at https://abcdstudy.org/federal-partners.html. A listing of participating sites and a complete listing of the study investigators can be found at https://abcdstudy.org/consortium_members/. ABCD Study consortium investigators designed and implemented the study and/or provided data but did not necessarily participate in analysis or writing of this report. This manuscript reflects the views of the authors and may not reflect the opinions or views of the NIH or ABCD Study consortium investigators. The ABCD Study data repository grows and changes over time. The ABCD data used in this report came from http://dx.doi.org/10.15154/cqdy-5453. Additional support for this work was made possible from NIEHS R01-ES032295 and R01-ES031074.

## Acknowledgments

The authors express acknowledgment and gratitude to the ABCD study participants and their families.

## Funding and assistance

This work is partially supported by the National Institutes of Health (NIH) National Institute of Diabetes and Digestive and Kidney Diseases R01 DK137899.

## Conflict of interest

The authors have nothing to disclose.

## Author contributions and guarantor statement

S.L. conceptualized this study. E.H. performed statistical analysis. A.C., E.H., and S.L. drafted the manuscript. All authors provided review, commentary, and revisions to the manuscript, and approved the final manuscript as submitted. S.L. is the guarantor of this work and, as such, had full access to all the data in the study and takes responsibility for the integrity of the data and the accuracy of the data analysis.

## Prior presentation

This work was presented as an oral presentation at the annual meeting of Western American Federation of Medical Research in 2024. During the course of preparing this work, the author(s) used ChatGPT (OpenAI, 2025) for the purpose of general editing and refining language. Following the use of this tool/service, the author(s) formally reviewed the content for its accuracy and edited it as necessary. The author(s) take full responsibility for all the content of this publication.

---

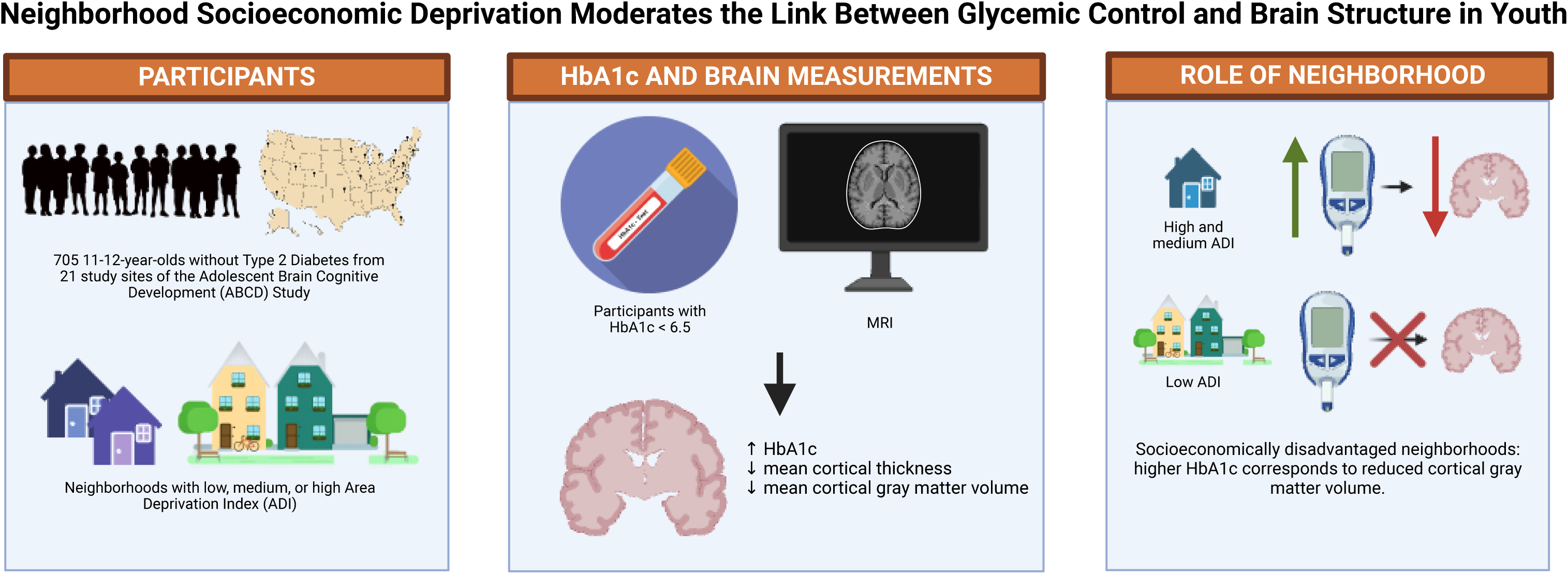

## Notes

### Competing Interest Statement

The authors have declared no competing interest.

### Author Declarations

Study procedures were approved by the centralized Institutional Review Board (IRB) at the University of California, San Diego (IRB #160091), in addition to each study site with its respective IRB. IRB of University of Southern California gave ethical approval for this work.

## References

1. National Diabetes Statistics Report | Diabetes | CDC. Accessed July 27, 2025. https://www.cdc.gov/diabetes/php/data-research/index.html

2. Lawrence JM, Divers J, Isom S, et al. Trends in Prevalence of Type 1 and Type 2 Diabetes in Children and Adolescents in the US, 2001-2017. JAMA. 2021;326(8):717. doi:10.1001/jama.2021.11165

3. Rodriquez IM, O’Sullivan KL. Youth-Onset Type 2 Diabetes: Burden of Complications and Socioeconomic Cost. Curr Diab Rep. 2023;23(5):59–67. doi:10.1007/s11892-023-01501-7

4. Nouwen A, Chambers A, Chechlacz M, et al. Microstructural abnormalities in white and gray matter in obese adolescents with and without type 2 diabetes. NeuroImage Clin. 2017;16:43–51. doi:10.1016/j.nicl.2017.07.004

5. Bruehl H, Sweat V, Tirsi A, Shah B, Convit A. Obese Adolescents with Type 2 Diabetes Mellitus Have Hippocampal and Frontal Lobe Volume Reductions. Neurosci Med. 2011;02(01):34–42. doi:10.4236/nm.2011.21005

6. Rofey DL, Arslanian SA, El Nokali NE, et al. Brain volume and white matter in youth with type 2 diabetes compared to obese and normal weight, nonldiabetic peers: A pilot study. Int J Dev Neurosci. 2015;46(1):88–91. doi:10.1016/j.ijdevneu.2015.07.003

7. Redel JM, DiFrancesco M, Vannest J, et al. Brain gray matter volume differences in obese youth with type 2 diabetes: a pilot study. J Pediatr Endocrinol Metab. 2018;31(3):261–268. doi:10.1515/jpem-2017-0349

8. Yau PL, Javier DC, Ryan CM, et al. Preliminary evidence for brain complications in obese adolescents with type 2 diabetes mellitus. Diabetologia. 2010;53(11):2298–2306. doi:10.1007/s00125-010-1857-y

9. Queen TL, Baucom KJW, Baker AC, Mello D, Berg CA, Wiebe DJ. Neighborhood disorder and glycemic control in late adolescents with Type 1 diabetes. Soc Sci Med. 2017;183:126–129. doi:10.1016/j.socscimed.2017.04.052

10. Khanolkar AR, Amin R, TaylorlRobinson D, Viner RM, Warner J, Stephenson T. Inequalities in glycemic control in childhood onset type 2 diabetes in England and Wales—A national populationlbased longitudinal study. Pediatr Diabetes. 2019;20(7):821–831. doi:10.1111/pedi.12897

11. Tremblay ES, Liu E, Laffel LM. Health Disparities Likely Emerge Early in the Course of Type-1 Diabetes in Youth. J Diabetes Sci Technol. 2022;16(4):929–933. doi:10.1177/19322968221082646

12. Garavan H, Bartsch H, Conway K, et al. Recruiting the ABCD sample: Design considerations and procedures. Dev Cogn Neurosci. 2018;32:16–22. doi:10.1016/j.dcn.2018.04.004

13. Yang R, Jernigan, Terry. Adolescent Brain Cognitive Development Study (ABCD) - Annual Release 4.0. doi:10.15154/1523041

14. Casey BJ, Cannonier T, Conley MI, et al. The Adolescent Brain Cognitive Development (ABCD) study: Imaging acquisition across 21 sites. Dev Cogn Neurosci. 2018;32:43–54. doi:10.1016/j.dcn.2018.03.001

15. Hagler DJ, Hatton S, Cornejo MD, et al. Image processing and analysis methods for the Adolescent Brain Cognitive Development Study. NeuroImage. 2019;202:116091. doi:10.1016/j.neuroimage.2019.116091

16. Fortin JP, Cullen N, Sheline YI, et al. Harmonization of cortical thickness measurements across scanners and sites. NeuroImage. 2018;167:104–120. doi:10.1016/j.neuroimage.2017.11.024

17. Fortin JP. neuroCombat: Harmonization of multi-site imaging data with ComBat. Published online 2023.

18. Singh GK. Area Deprivation and Widening Inequalities in US Mortality, 1969–1998. Am J Public Health. 2003;93(7):1137–1143. doi:10.2105/ajph.93.7.1137

19. Kind AJH, Jencks S, Brock J, et al. Neighborhood socioeconomic disadvantage and 30-day rehospitalization: a retrospective cohort study. Ann Intern Med. 2014;161(11):765–774. doi:10.7326/M13-2946

20. Mazaika PK, Weinzimer SA, Mauras N, et al. Variations in Brain Volume and Growth in Young Children With Type 1 Diabetes. Diabetes. 2016;65(2):476–485. doi:10.2337/db15-1242

21. Mauras N, Mazaika P, Buckingham B, et al. Longitudinal Assessment of Neuroanatomical and Cognitive Differences in Young Children With Type 1 Diabetes: Association With Hyperglycemia. Diabetes. 2015;64(5):1770–1779. doi:10.2337/db14-1445

22. Perantie DC, Wu J, Koller JM, et al. Regional brain volume differences associated with hyperglycemia and severe hypoglycemia in youth with type 1 diabetes. Diabetes Care. 2007;30(9):2331–2337. doi:10.2337/dc07-0351

23. Choi SE, Roy B, Freeby M, Mullur R, Woo MA, Kumar R. Prefrontal cortex brain damage and glycemic control in patients with type 2 diabetes. J Diabetes. 2020;12(6):465–473. doi:10.1111/1753-0407.13019

24. Canário N, Crisóstomo J, Duarte JV, et al. Irreversible atrophy in memory brain regions over 7 years is predicted by glycemic control in type 2 diabetes without mild cognitive impairment. Front Aging Neurosci. 2024;16:1367563. doi:10.3389/fnagi.2024.1367563

25. Zhang T, Shaw M, Cherbuin N. Association between Type 2 Diabetes Mellitus and Brain Atrophy: A Meta-Analysis. Diabetes Metab J. 2022;46(5):781–802. doi:10.4093/dmj.2021.0189

26. Vincent C, Hall PA. Executive Function in Adults With Type 2 Diabetes: A Meta-Analytic Review. Psychosom Med. 2015;77(6):631–642. doi:10.1097/PSY.0000000000000103

27. Garfield V, Farmaki AE, Eastwood SV, et al. HbA1c and brain health across the entire glycaemic spectrum. Diabetes Obes Metab. 2021;23(5):1140–1149. doi:10.1111/dom.14321

28. Coccaro EF, Lazarus S, Joseph J, et al. Emotional Regulation and Diabetes Distress in Adults With Type 1 and Type 2 Diabetes. Diabetes Care. 2021;44(1):20–25. doi:10.2337/dc20-1059

29. Roy B, Ehlert L, Mullur R, et al. Regional Brain Gray Matter Changes in Patients with Type 2 Diabetes Mellitus. Sci Rep. 2020;10(1):9925. doi:10.1038/s41598-020-67022-5

30. Wennberg AMV, Spira AP, Pettigrew C, et al. Blood glucose levels and cortical thinning in cognitively normal, middle-aged adults. J Neurol Sci. 2016;365:89–95. doi:10.1016/j.jns.2016.04.017

31. Zhang Y, Catts VS, Sheedy D, McCrossin T, Kril JJ, Shannon Weickert C. Cortical grey matter volume reduction in people with schizophrenia is associated with neuro-inflammation. Transl Psychiatry. 2016;6(12):e982. doi:10.1038/tp.2016.238

32. Patel Y, Parker N, Shin J, et al. Virtual Histology of Cortical Thickness and Shared Neurobiology in 6 Psychiatric Disorders. JAMA Psychiatry. 2021;78(1):47–63. doi:10.1001/jamapsychiatry.2020.2694

33. Li Q, Zhao Y, Chen Z, et al. Meta-analysis of cortical thickness abnormalities in medication-free patients with major depressive disorder. Neuropsychopharmacology. 2020;45(4):703–712. doi:10.1038/s41386-019-0563-9

34. Shin J, Patel Y, Parker N, Paus T, Pausova Z. Prediabetic HbA1c and Cortical Atrophy: Underlying Neurobiology. Diabetes Care. 2023;46(12):2267–2272. doi:10.2337/dc23-1105

