## Supplementary Materials for "The Role of Neighborhood Socioeconomic Environment in the Association Between Glycemic Control and the Developing Brain"

**Online-Only Supplemental Material**

**Supplemental Methods**

All covariate data was collected at two-year follow-up, with the following exceptions:

Sex at birth and race/ethnicity were collected at baseline. Baseline Area Deprivation Index (ADI) was imputed for missing 2-year follow-up ADI, when available, as ADI was highly correlated between the two time points (r=0.995, *P*<0.001), and did not change significantly over time (β (95% CI) = 0.073(-0.01,0.16), P=0.090).

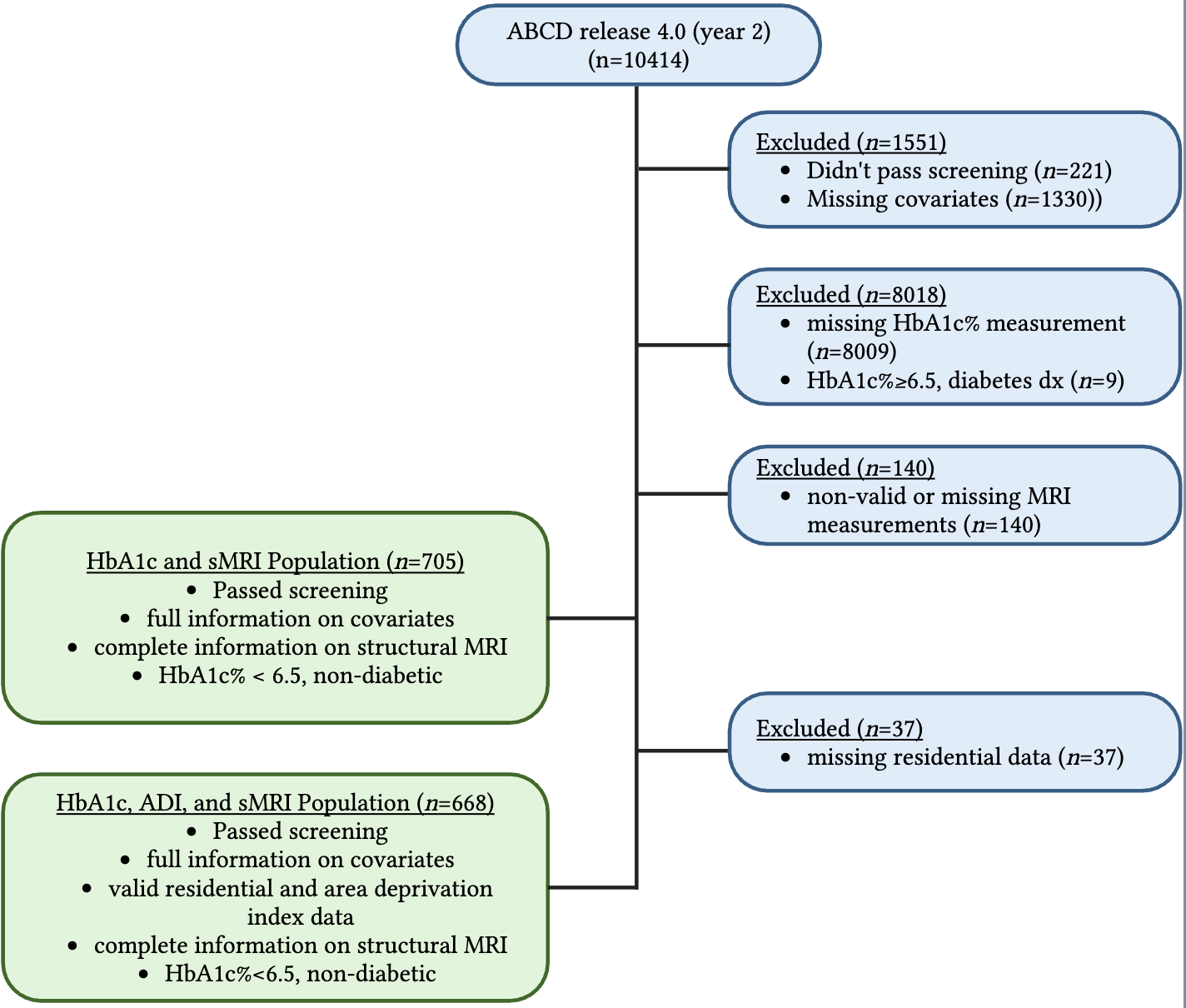
Supplementary Figure 1. Sample flowchart depicts exclusions of participants based on 1) screening for eligibility and medical history, and missing data for study covariates (age, sex, race/ethnicity, family income, family education, and pubertal stage, 2) non-availability of HbA1c measurement and positive diabetes diagnosis status, 3) non-validity and non-availability of MRI data, and 4) non-availability of residential data. A sample of n = 705 was used for main analysis of association between HbA1c and sMRI, and a reduced sample of n = 668 was used for sub-analysis of association between HbA1c and sMRI modulated by ADI.

**Table S1. Associations between HbA1c and Regional Cortical Gray Matter Volume**

| **Regional Cortical Gray Matter Volume (mm^3^)** | **β (95% CI)*** | **FDR-adjusted *P*-value†** |
| --- | --- | --- |
| Banks of the Superior Temporal Sulcus | 0.014 (-0.038, 0.066) | 0.736 |
| Caudal Anterior Cingulate Cortex | -0.016 (-0.065, 0.033) | 0.736 |
| Caudal Middle Frontal Gyrus | -0.032 (-0.089, 0.025) | 0.505 |
| Cuneus Cortex | -0.075 (-0.138, -0.011) | 0.102 |
| Entorhinal Cortex | -0.016 (-0.073, 0.041) | 0.736 |
| Fusiform Gyrus | -0.016 (-0.064, 0.033) | 0.736 |
| Inferior Parietal Cortex | 0.014 (-0.038, 0.066) | 0.736 |
| Inferior Temporal Gyrus | -0.052 (-0.102, -0.002) | 0.176 |
| Isthmus Cingulate Cortex | 0.026 (-0.026, 0.079) | 0.581 |
| Lateral Occipital Cortex | -0.093 (-0.148, -0.039) | **0.025** |
| Lateral Orbital Frontal Cortex | -0.047 (-0.097, 0.003) | 0.253 |
| Lingual Gyrus | -0.045 (-0.104, 0.014) | 0.420 |
| Medial Orbital Frontal Cortex | 0.003 (-0.044, 0.05) | 0.900 |
| Middle Temporal Gyrus | -0.017 (-0.069, 0.034) | 0.736 |
| Parahippocampal Gyrus | -0.083 (-0.142, -0.023) | 0.053 |
| Paracentral Lobule | -0.079 (-0.132, -0.026) | **0.040** |
| Pars Opercularis | -0.008 (-0.065, 0.050) | 0.892 |
| Pars Orbitalis | -0.072 (-0.126, -0.017) | 0.060 |
| Pars Triangularis | -0.044 (-0.103, 0.014) | 0.420 |
| Pericalcarine Cortex | -0.045 (-0.112, 0.023) | 0.438 |
| Postcentral Gyrus | -0.030 (-0.082, 0.022) | 0.502 |
| Posterior Cingulate Cortex | -0.065 (-0.114, -0.017) | 0.059 |
| Precentral Gyrus | -0.024 (-0.073, 0.025) | 0.584 |
| Precuneus Cortex | -0.006 (-0.057, 0.044) | 0.892 |
| Rostral Anterior Cingulate Cortex | -0.004 (-0.053, 0.046) | 0.900 |
| Rostral Middle Frontal Gyrus | -0.011 (-0.064, 0.042) | 0.789 |
| Superior Frontal Gyrus | -0.078 (-0.127, -0.030) | **0.029** |
| Superior Parietal Cortex | 0.021 (-0.032, 0.074) | 0.712 |
| Superior Temporal Gyrus | -0.037 (-0.092, 0.018) | 0.438 |
| Supramarginal Gyrus | 0.016 (-0.037, 0.069) | 0.736 |
| Frontal Pole | 0.004 (-0.051, 0.059) | 0.900 |
| Temporal Pole | -0.042 (-0.102, 0.018) | 0.432 |
| Transverse Temporal Cortex | -0.035 (-0.095, 0.025) | 0.502 |
| Insular Cortex | -0.037 (-0.09, 0.015) | 0.432 |

* Standardized regression coefficient (95% CI) from linear mixed effects models. For regional cortical gray matter volumes, coefficient (95% CI) represents main effect estimate of HbA1c. HbA1c by hemisphere interactions were modeled but not significant in any region of interest.

† Multiple comparisons are conducted with Benjamini-Hochberg FDR correction. Boldface indicates significance at the corrected threshold of *P*<0.05.

**Table S2. Associations between HbA1c and Regional Cortical Thickness**

| **Regional Cortical Thickness (mm)** | **β (95% CI)*** | **FDR-adjusted *P*-value†** |
| --- | --- | --- |
| Banks of the Superior Temporal Sulcus | -0.036 (-0.098, 0.026) | 0.308 |
| Caudal Anterior Cingulate Cortex | -0.079 (-0.140, -0.017) | 0.070 |
| Caudal Middle Frontal Gyrus | 0.002 (-0.066, 0.070) | 0.949 |
| Cuneus Cortex | -0.068 (-0.134, -0.001) | 0.100 |
| Entorhinal Cortex | 0.007 (-0.058, 0.072) | 0.851 |
| Fusiform Gyrus | -0.04 (-0.107, 0.026) | 0.308 |
| Inferior Parietal Cortex | -0.071 (-0.139, -0.003) | 0.100 |
| Inferior Temporal Gyrus | -0.087 (-0.154, -0.019) | 0.070 |
| Isthmus Cingulate Cortex | -0.107 (-0.172, -0.043) | **0.018** |
| Lateral Occipital Cortex | -0.101 (-0.170, -0.033) | **0.037** |
| Lateral Orbital Frontal Cortex | -0.067 (-0.133, -0.001) | 0.100 |
| Lingual Gyrus | -0.071 (-0.139, -0.004) | 0.100 |
| Medial Orbital Frontal Cortex | -0.047 (-0.109, 0.015) | 0.213 |
| Middle Temporal Gyrus | -0.08 (-0.147, -0.013) | 0.095 |
| Parahippocampal Gyrus | -0.059 (-0.123, 0.006) | 0.139 |
| Paracentral Lobule | -0.075 (-0.141, -0.009) | 0.100 |
| Pars Opercularis | -0.033 (-0.098, 0.031) | 0.362 |
| Pars Orbitalis | -0.093 (-0.157, -0.029) | **0.037** |
| Pars Triangularis | -0.055 (-0.121, 0.011) | 0.174 |
| Pericalcarine Cortex | -0.041 (-0.110, 0.027) | 0.308 |
| Postcentral Gyrus | -0.062 (-0.129, 0.005) | 0.136 |
| Posterior Cingulate Cortex | -0.107 (-0.171, -0.044) | **0.018** |
| Precentral Gyrus | -0.019 (-0.086, 0.048) | 0.616 |
| Precuneus Cortex | -0.078 (-0.146, -0.010) | 0.100 |
| Rostral Anterior Cingulate Cortex | -0.062 (-0.123, -0.002) | 0.100 |
| Rostral Middle Frontal Gyrus | -0.035 (-0.104, 0.034) | 0.369 |
| Superior Frontal Gyrus | -0.05 (-0.122, 0.021) | 0.238 |
| Superior Parietal Cortex | -0.041 (-0.112, 0.029) | 0.308 |
| Superior Temporal Gyrus | -0.071 (-0.141, -0.002) | 0.100 |
| Supramarginal Gyrus | -0.048 (-0.116, 0.02) | 0.238 |
| Frontal Pole | -0.019 (-0.082, 0.045) | 0.616 |
| Temporal Pole | -0.065 (-0.129, -0.002) | 0.100 |
| Transverse Temporal Cortex | -0.056 (-0.119, 0.007) | 0.145 |
| Insular Cortex | -0.052 (-0.116, 0.011) | 0.174 |

* Standardized regression coefficient (95% CI) from linear mixed effects models. For regional cortical thickness measurements, coefficient (95% CI) represents main effect estimate of HbA1c. HbA1c by hemisphere interactions were modeled but not significant in any region of interest.

† Multiple comparisons are conducted with Benjamini-Hochberg FDR correction. Boldface indicates significance at the corrected threshold of *P*<0.05.

**Table S3. Associations between HbA1c and Regional Cortical Gray Matter Volume, by Area Deprivation Index (ADI)**

|  | **High ADI** | | **Medium ADI** | | **Low ADI** | |
| --- | --- | --- | --- | --- | --- | --- |
| **Regional Cortical Gray Matter Volume (mm^3^)** | **β (95% CI)*** | **FDR-adjusted *P*-value****†** | **β (95% CI)*** | **FDR-adjusted *P*-value†** | **β (95% CI)*** | **FDR-adjusted *P*-value†** |
| Lateral Occipital Cortex | -0.093 (-0.182, -0.004) | **0.040** | -0.145 (-0.248, -0.041) | **0.019** | -0.074 (-0.164, 0.017) | 0.328 |
| Paracentral Lobule | -0.111 (-0.199, -0.023) | **0.020** | -0.083 (-0.185, 0.019) | 0.128 | -0.046 (-0.136, 0.043) | 0.458 |
| Superior Frontal Cortex | -0.133 (-0.215, -0.052) | **0.004** | -0.073 (-0.168, 0.021) | 0.128 | -0.031 (-0.114, 0.052) | 0.458 |

* Standardized regression coefficient (95% CI) from linear mixed effects models. For regional cortical gray matter volumes measurements, coefficient (95% CI) represents least-squares estimates of HbA1c within ADI tertile. HbA1c by hemisphere interactions were modeled but not significant in any region of interest.

† Multiple comparisons are conducted with Benjamini-Hochberg FDR correction. Boldface indicates significance at the corrected threshold of *P*<0.05.

**Table S4. Associations between HbA1c and Regional Cortical Thickness, by Area Deprivation Index**

|  | **High ADI** | | **Medium ADI** | | **Low ADI** | |
| --- | --- | --- | --- | --- | --- | --- |
| **Regional Cortical Thickness (mm)** | **β (95% CI)*** | **FDR-adjusted *P*-value†** | **β (95% CI)*** | **FDR-adjusted *P*-value†** | **β (95% CI)*** | **FDR-adjusted *P*-value†** |
| Isthmus Cingulate Cortex | -0.071 (-0.177, 0.036) | 0.193 | -0.149 (-0.274, -0.024) | **0.038** | -0.141 (-0.250, -0.032) | **0.045** |
| Lateral Occipital Cortex | -0.162 (-0.275, -0.050) | **0.011** | -0.132 (-0.263, -0.001) | 0.065 | -0.013 (-0.128, 0.102) | 0.820 |
| Pars Orbitalis | -0.150 (-0.255, -0.045) | **0.011** | -0.090 (-0.212, 0.033) | 0.151 | -0.049 (-0.157, 0.058) | 0.600 |
| Posterior Cingulate Cortex | -0.131 (-0.235, -0.027) | **0.018** | -0.188 (-0.309, -0.066) | **0.010** | -0.041 (-0.147, 0.065) | 0.600 |

* Standardized regression coefficient (95% CI) from linear mixed effects models. For regional cortical thickness measurements, coefficient (95% CI) represents least-squares estimates of HbA1c within ADI tertile. HbA1c by hemisphere interactions were modeled but not significant in any region of interest.

† Multiple comparisons are conducted with Benjamini-Hochberg FDR correction. Boldface indicates significance at the corrected threshold of *P*<0.05.
